# Prognosis Analysis of Liver Transplantation for Hepatocellular Carcinoma and Intrahepatic Cholangiocarcinoma in Stage 6 AJCC I: A SEER Database Study

**DOI:** 10.64898/2026.01.30.26345191

**Authors:** Chuan Jiang, Linhui Gong

## Abstract

**Background:** Liver transplantation (LT) offers a curative option for early-stage hepatocellular carcinoma (HCC). Its role in intrahepatic cholangiocarcinoma (ICC) remains controversial, with limited comparative evidence on long-term outcomes, especially for early-stage disease.

**Methods:** This retrospective population-based study utilized the SEER database (2004-2015). Patients with AJCC 6th edition Stage I HCC or ICC who underwent LT were included. Cancer-specific survival (CSS) and overall survival (OS) were primary endpoints. Kaplan-Meier ana lysis, log-rank tests, and Cox proportional hazards regression were used for survival comparison and identification of prognostic factors.

**Results:** Among 944 eligible patients, 925 had HCC and 19 had ICC. The 5-year OS and CSS rates were significantly higher for HCC patients (OS: 95.1%; CSS: 97.7%) compared to ICC patients (OS: 82.3%; CSS: 82.3%). Multivariate Cox analysis for HCC identified age and marital status as independent risk factors for OS, and tumor size for CSS. For ICC, only tumor size was associated with OS in univariate analysis; no independent risk factors for CSS were identified due to the small sample size.

**Conclusions:** LT provides excellent long-term survival for patients with early-stage HCC. In contrast, outcomes for early-stage ICC patients after LT are significantly inferior. Prognostic factors differ between the two histological types, underscoring the need for distinct LT selection criteria and management strategies. The findings highlight the limited utility of LT for ICC based on current selection paradigms and emphasize the necessity for larger studies incorporating molecular pro filing to identify potential ICC subpopulations that may benefit from LT.

## Introduction

Primary liver cancer ranks as the sixth most common malignant tumor globally and the fourth leading cause of cancer-related deaths. Its incidence continues to rise, posing a severe challenge to public health systems worldwide[1]. HCC and ICC constitute the predominant subtypes, accounting for approximately 75% and 15% of all cases, respectively[2]. Hepatic resection remains the preferred treatment for early-stage primary liver cancer, particularly in patients with localized tumors and adequate liver reserve[3–5]. However, its feasibility is often limited by the severity of underlying liver disease and insufficient residual liver volume post-surgery, and it fails to address the underlying liver pathology at its root[6]. Therefore, for patients with early-stage primary liver cancer complicated by chronic liver diseases such as cirrhosis, LT demonstrates irreplaceable value—it not only achieves negative oncological margins and eradicates intrahepatic micrometastases but also completely resolves the underlying liver disease[7–9]. However, the applicability of liver transplantation in ICC treatment remains highly controversial. Most studies suggest that ICC exhibits greater invasiveness and a tendency toward early micrometastasis, often rendering it unsuitable for liver transplantation[10–12]. While transplant oncology concepts have expanded indications for liver transplantation, its efficacy for early-stage ICC patients remains unclear[13–14].

Current studies comparing liver transplantation outcomes between H CC and ICC are predominantly single-center retrospective or small-sample analyses, yielding limited evidence. There is a particular lack of large-population, long-term comparative studies focusing on early-stage (AJCC Stage I) patients, hindering our understanding of the true efficacy of liver transplantation for both primary liver cancers at an early tumor stage. The National Cancer Institute’s Surveillance, Epidemiology, and End Results (SEER) database, covering approximately 28% of the U.S. population, provides a platform for conducting large-scale, long-term real-world studies.

This study represents the first systematic comparison using the SEE R database to evaluate long-term survival outcomes in HCC and IC C patients who underwent liver transplantation and met the AJCC 6th edition Stage I criteria. Using CSS and OS as primary endpoints, it assesses prognostic risk factors and differences in treatment efficacy between early-stage HCC and ICC following liver transplantation.

## Materials and methods

### Patient selection

Based on the US SEER database, patients diagnosed with HCC and ICC between 2004 and 2015 were selected. ICC corresponds to primary site code C22.1 and ICD-O-3 histology code 8160/3; HCC corresponded to C22.1 with an ICD-O-3 code of 8170/3. The study included patients who underwent LT (Surg Prim Site=61) and collected data on age, race, marital status, tumor size, receipt of chemotherapy and radiotherapy, number of lymph nodes removed, and survival information. Concurrently, we excluded cases involving patients younger than 18 years old, as well as those with missing data on race, marital status, tumor size, chemotherapy, radiotherapy, number of lymph nodes removed, or survival status. Since the SEER database provides publicly accessible anonymized data, no IRB (Institutional Review Board) approval or explicit consent was required.

### Statistical analysis

The primary endpoints were CSS and OS. CSS was defined as the time (in months) from HCC or ICC diagnosis to follow-up cutoff or death from HCC or ICC. Patients lost to follow-up and those who died from causes other than HCC or ICC were treated as missing data. OS was defined as the time (in months) from diagnosis to follow-up completion or death from any cause, with patients lost to follow-up treated as missing data. Data description and intergroup comparisons: Normality of quantitative data was assessed using the Kolmogorov-Smirnov test. Normally distributed quantitative data were expressed as mean ± standard deviation (Mean ± SD), while skewed quantitative data were presented as median and interquartile range (Median (IQR)). Intergroup comparisons were performed using the Mann-Whitney U test. Categorical data were expressed as frequency and percentage. Intergroup comparisons were performed using the chi-square (χ²) test or Fisher’s exact test. For survival analysis and prognostic factor evaluation: Continuous variables were converted to binary variables using X-tile 3.6.1 software to determine optimal cutoff points. Univariate Cox proportional hazards regression models analyzed the relationship between each variable and CSS or OS in HCC or ICC patients. Variables showing statistical significance (P < 0.05) in univariate analysis were included in multivariate Cox proportional hazards regression models. Multivariate Cox regression analysis identified independent prognostic factors associated with CSS or OS in HCC or ICC patients, calculating their hazard ratios (HR) and corresponding 95% confidence intervals (95% CI). Survival curves were plotted using the Kaplan-Meier method, and intergroup survival differences were compared using the log-rank test. All hypothesis tests were two-sided with a significance level of α = 0.05. Optimal cutoff values were determined using X-tile 3.6.1 software. Statistical analysis, Cox regression analysis, and survival analysis were performed using SPSS 22.0 and R 4.5.1 software.

## Results

### Patients characteristics

A total of 944 patients were included, comprising 925 in the HCC group and 19 in the ICC group. The Kolmogorov-Smirnov test indicated that tumor size and age in both groups did not follow a nor mal distribution (P < 0.05). Data are expressed as median (interquartile range): tumor size was 22 (15–32) mm in the HCC group and 20 (15–35) mm in the ICC group; age was 57 (53–62) years in the HCC group and 60 (53–64) years in the ICC group. No statistically significant differences were observed between groups in baseline characteristics (Table 1).

**Table 1.** Baseline characteristics of HCC and ICC patients.

| Variable factors | HCC (n=925) | ICC (n=19) | P value |
| --- | --- | --- | --- |
| Age(Years) |  |  |  |
| Median | 57 | 60 | 0.334 |
| IQR | 53-62 | 53-64 |  |
| Sex |  |  |  |
| Male | 680(73.5%) | 12(63.1%) | 0.312 |
| Female | 245(26.5%) | 7(36.9%) |  |
| Race |  |  |  |
| White | 744(80.4%) | 16(84.2%) | 0.725 |
| Black | 85(9.2%) | 2(10.5%) |  |
| Other | 96(10.4) | 1(5.3%) |  |
| Tumor size(mm) |  |  |  |
| Median | 22 | 20 | 0.864 |
| IQR | 15-32 | 15-35 |  |
| Marriage |  |  |  |
| Married | 624(67.5%) | 13(68.4%) | 0.929 |
| Unmarried | 301(32.5%) | 6(31.6%) |  |
| Chemotherapy |  |  |  |
| Done | 281(30.4%) | 7(36.8%) | 0.545 |
| None | 644(69.6%) | 12(63.2%) |  |
| Radiation |  |  |  |
| Done | 15(1.6%) | 0(0%) | 1 |
| None | 910(98.4%) | 19(100%) |  |
| Lymph |  |  |  |
| Done | 665(71.9%) | 12(63.1%) | 0.682 |
| 1-3piece | 233(25.2%) | 6(31.6%) |  |
| >=4piece | 27(2.9%) | 1(5.3%) |  |

Values were presented n (%) or median (IQR)

### Survival analysis

Results showed that the 1-year, 3-year, and 5-year OS rates for patients with HCC undergoing liver transplantation were 99.7%, 97.9%, and 95.1%, respectively, with corresponding CSS rates of 99.2%, 99.1%, and 97.7%. In contrast, among patients with ICC who underwent liver transplantation, the 1-year, 3-year, and 5-year OS rates were 97.8%, 94.7%, and 82.3%, respectively, while the CSS rates were 97.8%, 94.7%, and 82.3%. Comparisons reveal that both OS and CSS rates were lower in ICC patients than in HCC patients (Table 2). Additionally, survival curves illustrate the trends in postoperative OS and CSS for both patient groups (Figure 1 and Figure 2).

**Fig 1.**
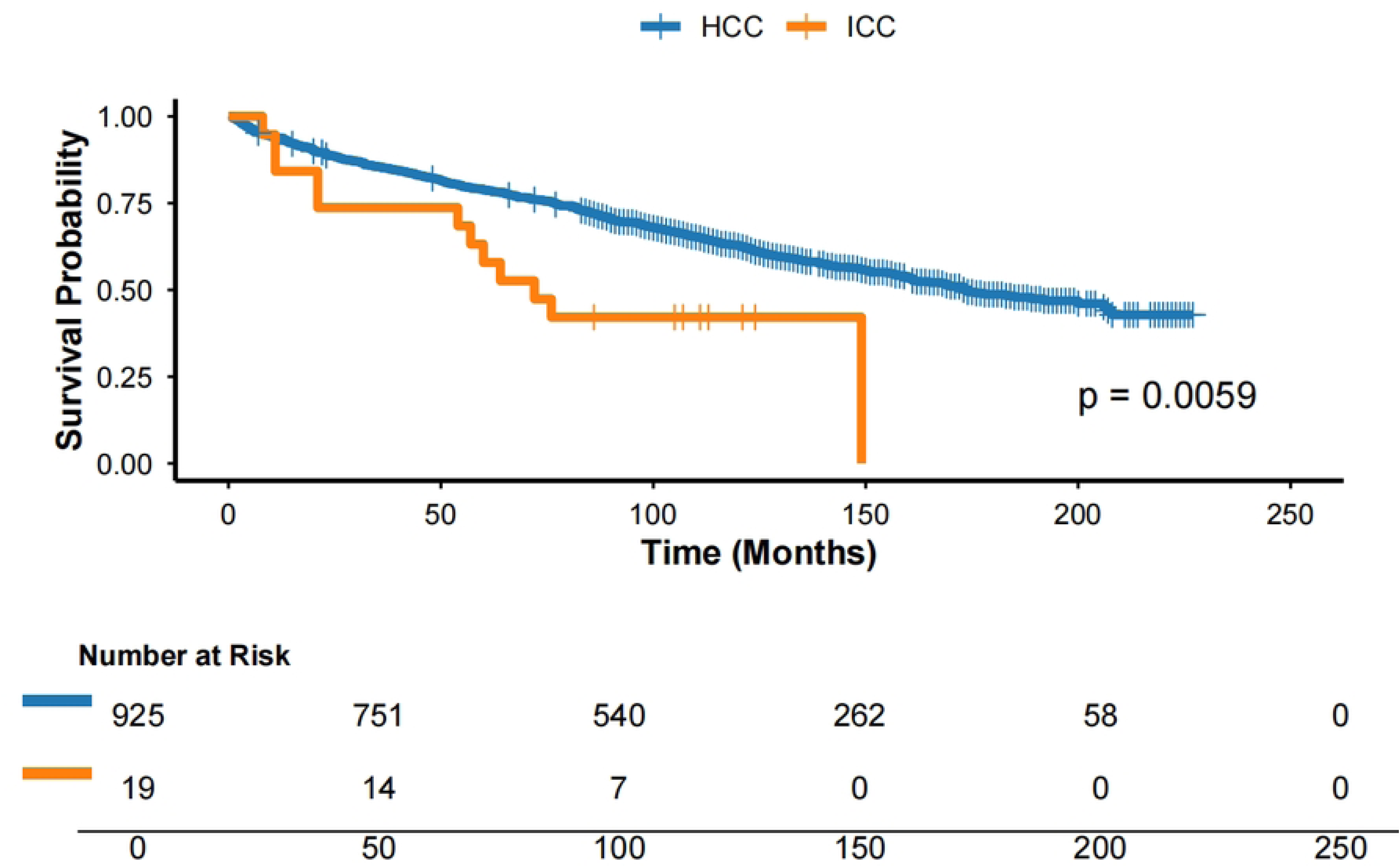
CSS curves for two groups after LT.

**Fig 2.**
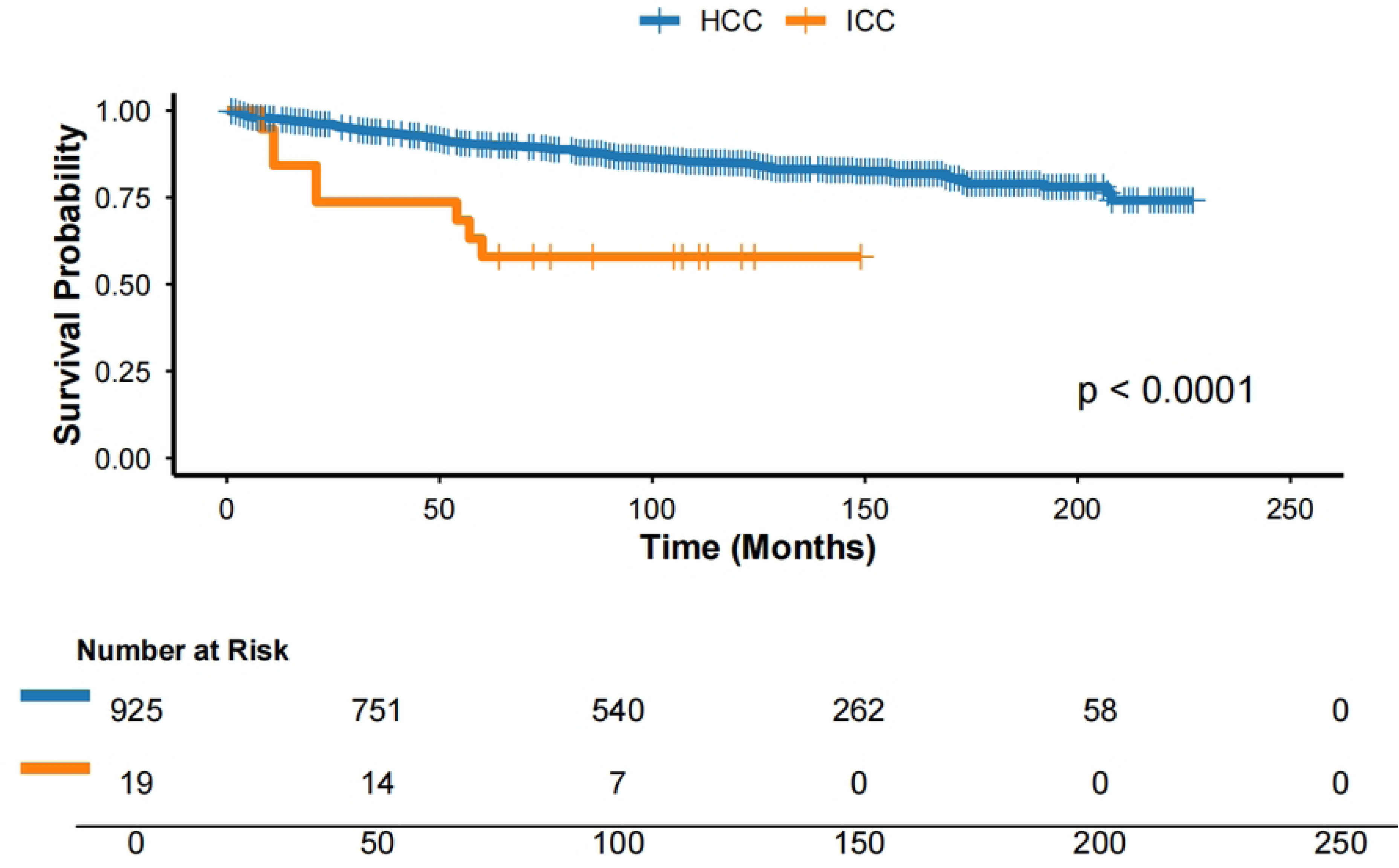
OS curves for two groups after LT.

**Table 2.** Survival outcomes of patients with HCC and ICC.

| Outcome | HCC(n=925) | ICC(n=19) | P value |
| --- | --- | --- | --- |
| Overall survival |  |  |  |
| 1years | 99.7% | 97.8% | <0.001 |
| 3years | 97.9% | 94.7% | <0.001 |
| 5years | 95.1% | 82.3% | <0.001 |
| Cancer-specific survival |  |  |  |
| 1years | 99.2% | 97.8% | <0.001 |
| 3years | 99.1% | 94.7% | <0.001 |
| 5years | 97.7% | 82.3% | <0.001 |

Values were presented n (%)

### Univariate and multivariate analysis of HCC

Among 925 HCC patients who underwent LT, the optimal cutoff values for continuous variables were determined using X-tile software and converted into binary variables: age was stratified at 63 years for OS analysis (Figure 3) and at 64 years for CSS analysis (Figure 4); tumor size was stratified at 43 mm for OS analysis (Figure 5) and at 43 mm for CSS analysis (Figure 6). In univariate Cox regression analysis, age and marital status were significant predictors of OS. Multivariate analysis further confirmed both as independent risk factors for OS (Table 3). Regarding CSS, univariate analysis suggested tumor size and age were associated with CSS, while multivariate analysis identified tumor size as an independent risk factor for CSS (Table 4).

**Fig 3.**
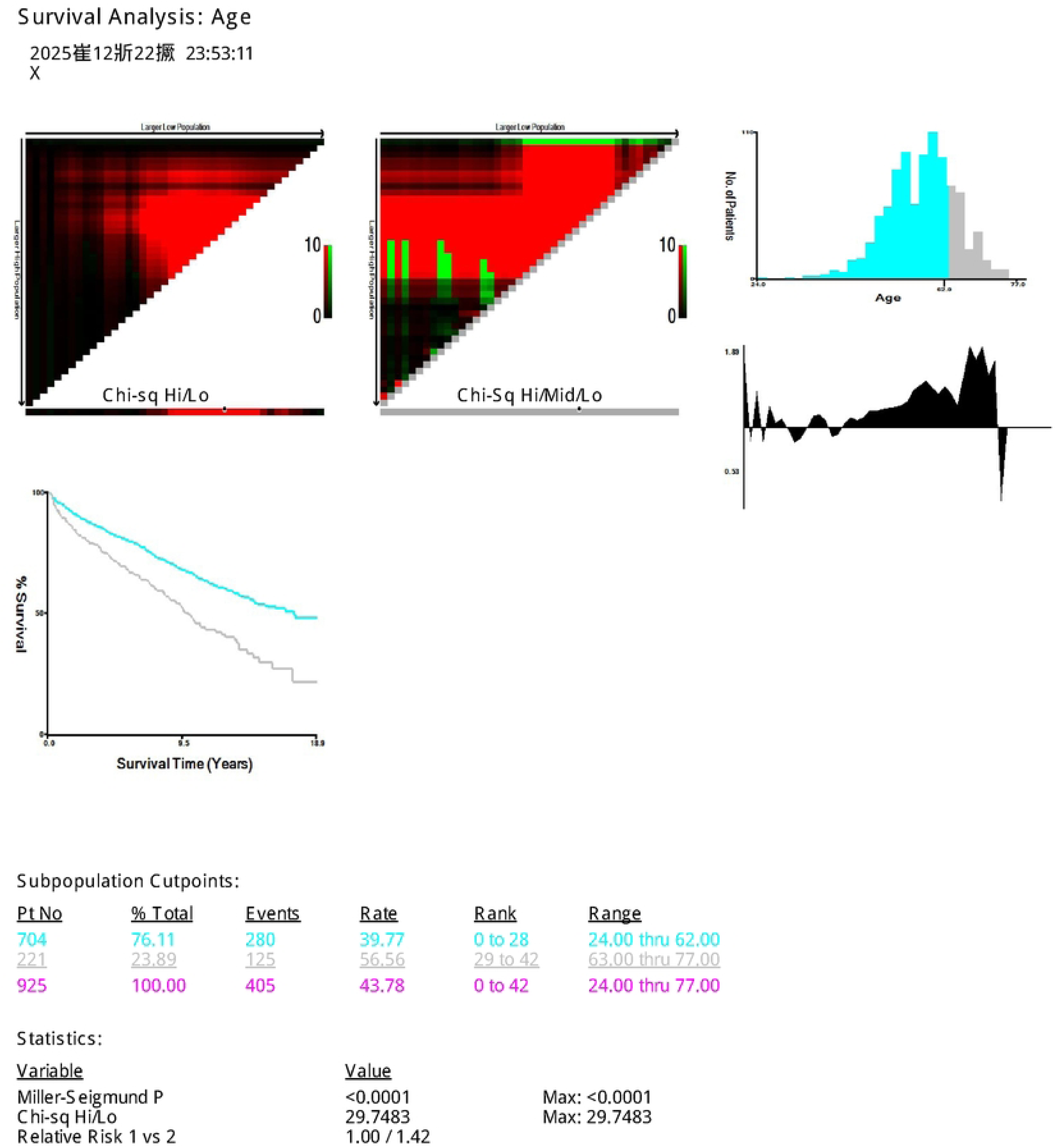
Optimal cutoff age for OS in HCC.

**Fig 4.**
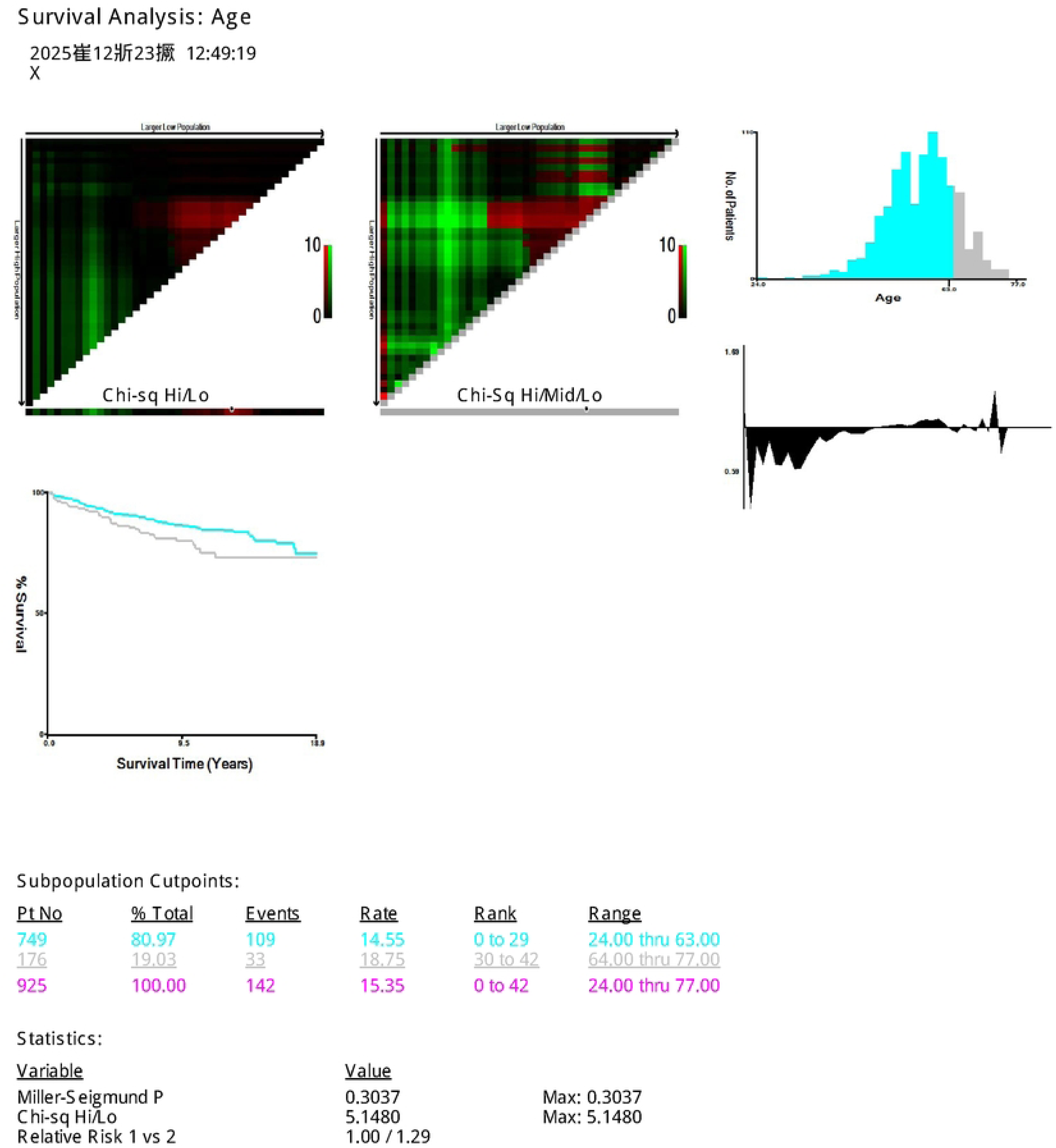
Optimal cutoff age for CSS in HCC.

**Fig 5.**
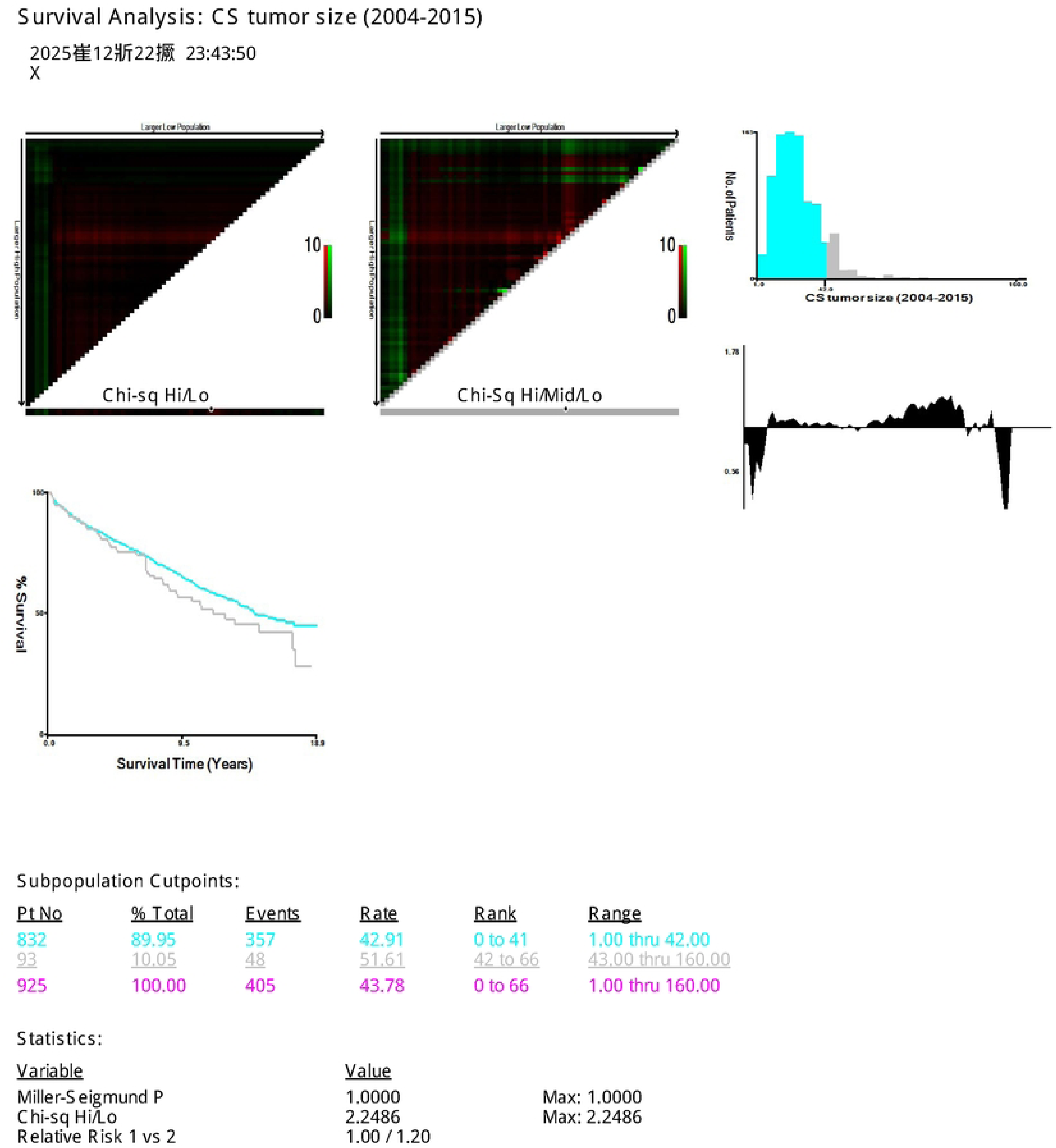
Optimal cutoff tumor size for OS in HCC.

**Fig 6.**
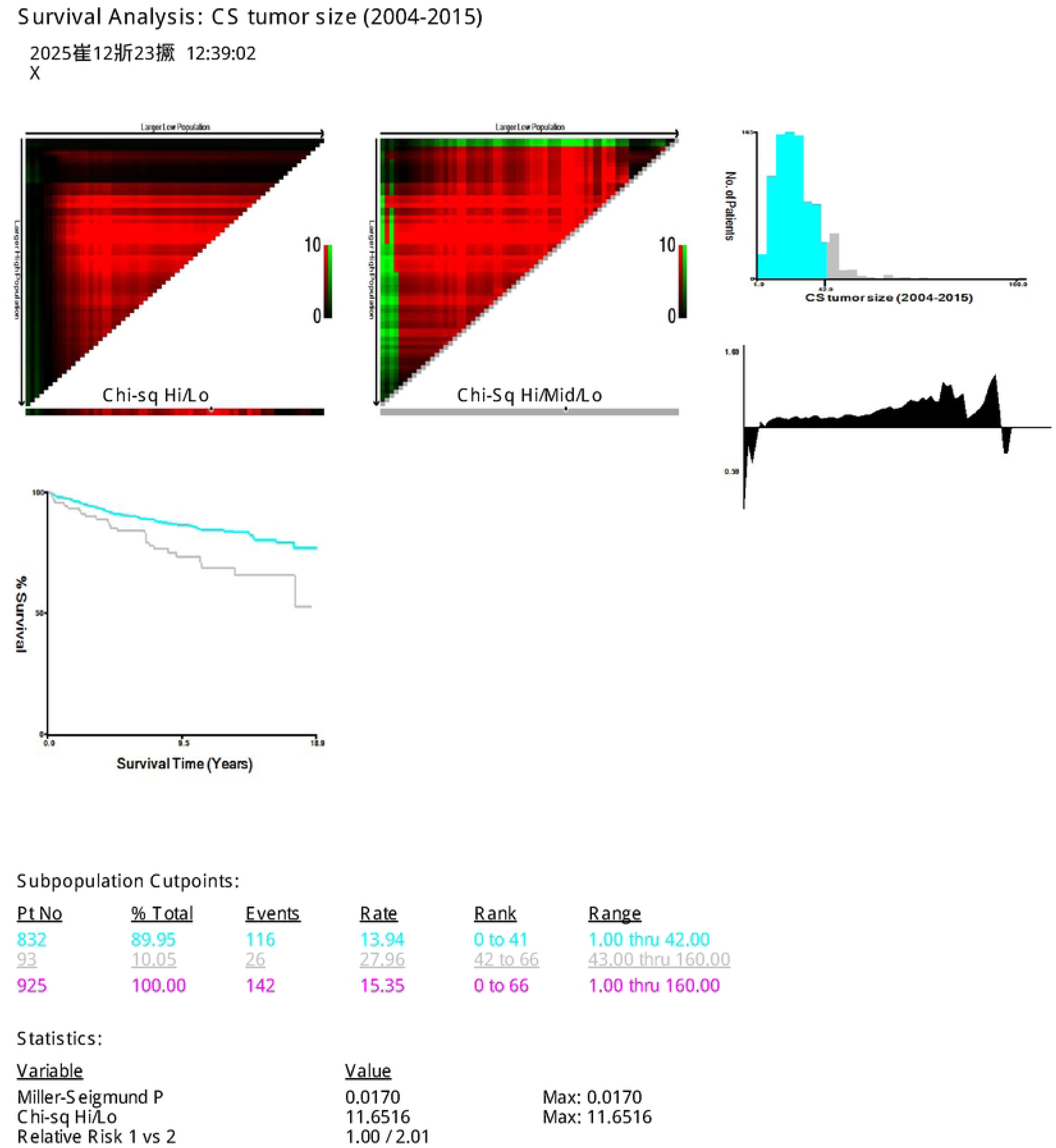
Optimal cutoff tumor size for CSS in HCC.

**Table 3.** Univariate and Multivariate analyses for Overall survival in HCC patients.

| Variable factors | Overall survival |  | Multivariate analysis |  |
| --- | --- | --- | --- | --- |
|  | Univariate analysis<br>HR (95% CI) | P<br>value | HR (95% CI) | P<br>value |
| Age |  |  |  |  |
| >=63years | Ref |  | Ref |  |
| <63years | 0.559(0.452-0.691) | 0.001 | 0.557(0.451-0.689) | 0.001 |
| Sex |  |  |  |  |
| Female | Ref |  |  |  |
| Male | 0.948(0.785-1.185) | 0.639 |  |  |
| Race |  |  |  |  |
| Other | Ref |  |  |  |
| White | 1.161(0.826-1.631) | 0.391 |  |  |
| Black | 1.510(0.967-2.356) | 0.070 |  |  |
| Marriage |  |  |  |  |
| Unmarried | Ref |  | Ref |  |
| Married | 0.799(0.652-0.980) | 0.032 | 1.260(1.027-1.545) | 0.027 |
| Chemotherapy |  |  |  |  |
| None | Ref |  |  |  |
| Done | 3.079(0.923-10.273) | 0.067 |  |  |
| Radiation |  |  |  |  |
| None | Ref |  |  |  |
| Done | 1.161(0.576-2.337) | 0.677 |  |  |
| Tumor size |  |  |  |  |
| >=43mm | Ref |  |  |  |
| <43mm | 0.793(0.587-1.072) | 0.132 |  |  |
| Lymph node |  |  |  |  |
| >=4piece | Ref |  |  |  |
| None | 1.147(0.628-2.095)<sup>10</sup> | 0.655 |  |  |
| 1-3piece | 1.218(0.654-2.267) | 0.534 |  |  |

Abbreviations: HR, hazard ratio; CI, confidence interval.

**Table 4.**
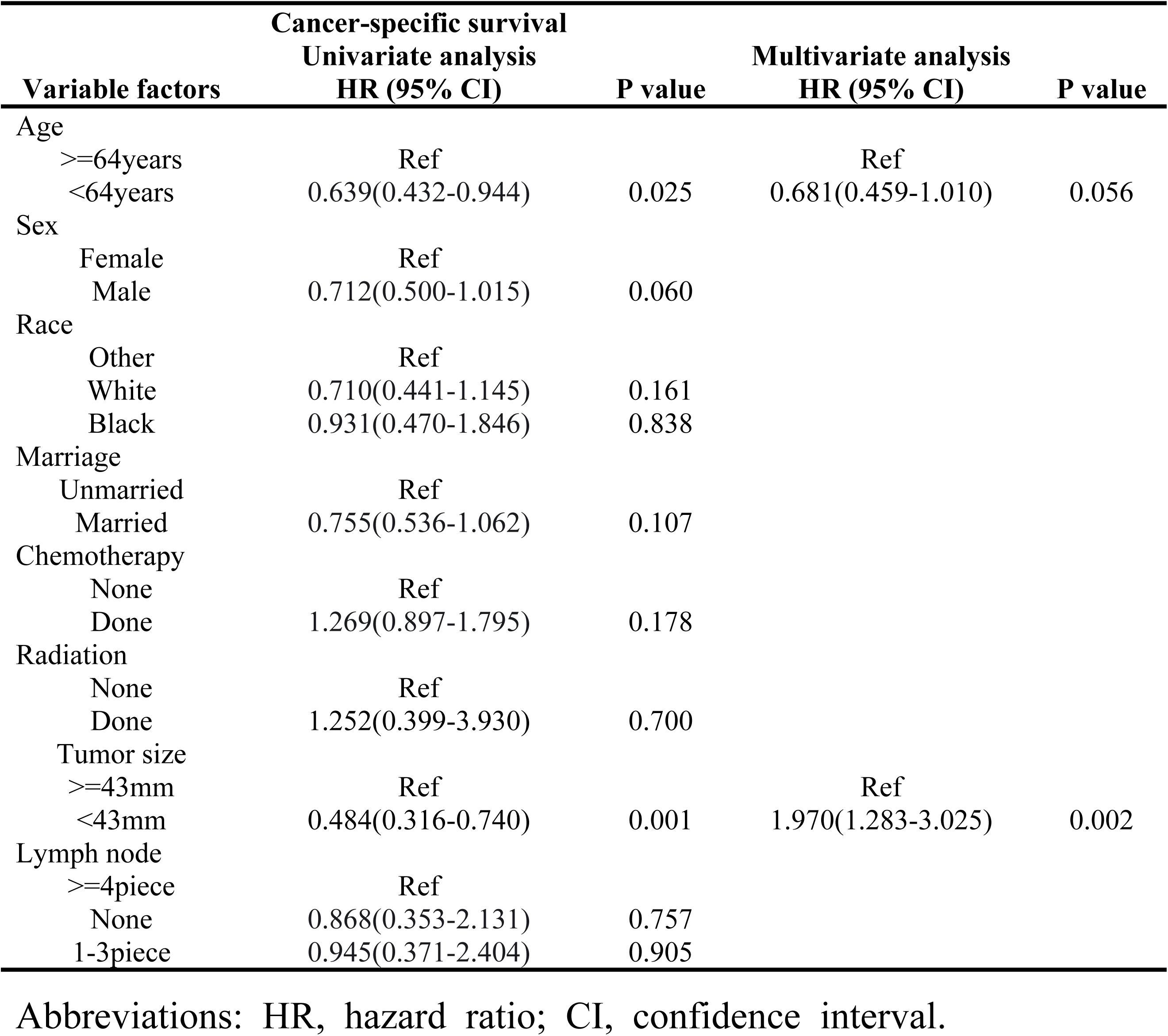
Univariate and Multivariate analyses for Cancer-specific survival in HCC patients.

### Univariate and multivariate analysis of ICC

Among 19 patients with ICC who underwent LT, the continuous variables age and tumor size were converted into categorical variables using optimal cutoff points determined by X-tile software: age was categorized at 60 years for OS analysis (Figure 7) and at 47 years for CSS analysis (Figure 8); Tumor size was categorized at 29 mm for OS analysis (Figure 9) and at 13 mm for CSS analysis (Figure 10). Univariate Cox regression analysis revealed that tumor size was significantly associated with OS(Table 5). However, no statistically significant risk factors were identified for CSS(Table 6).

**Fig 7.**
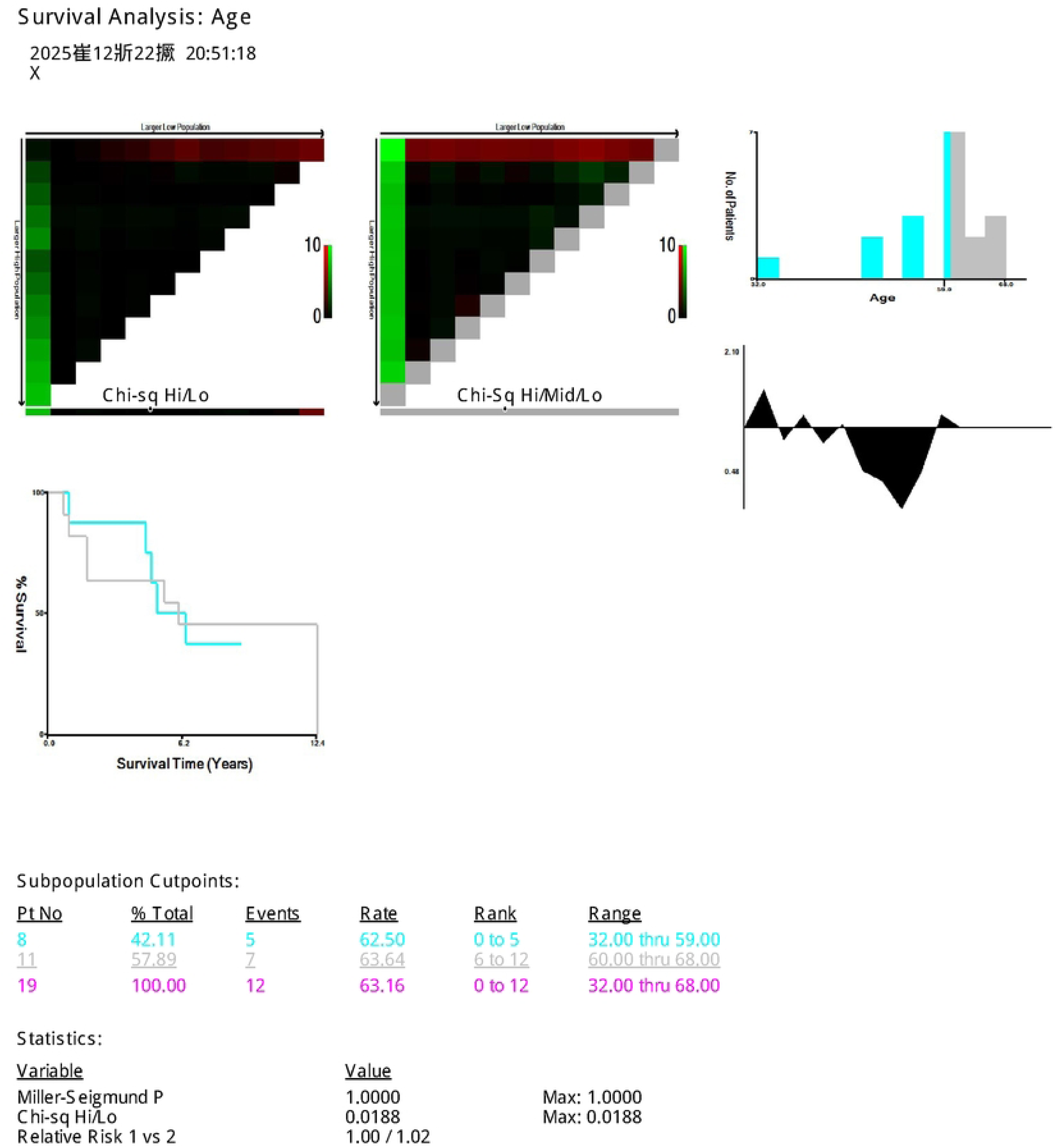
Overall survival curves for two groups after liver transplantation.

**Fig 8.**
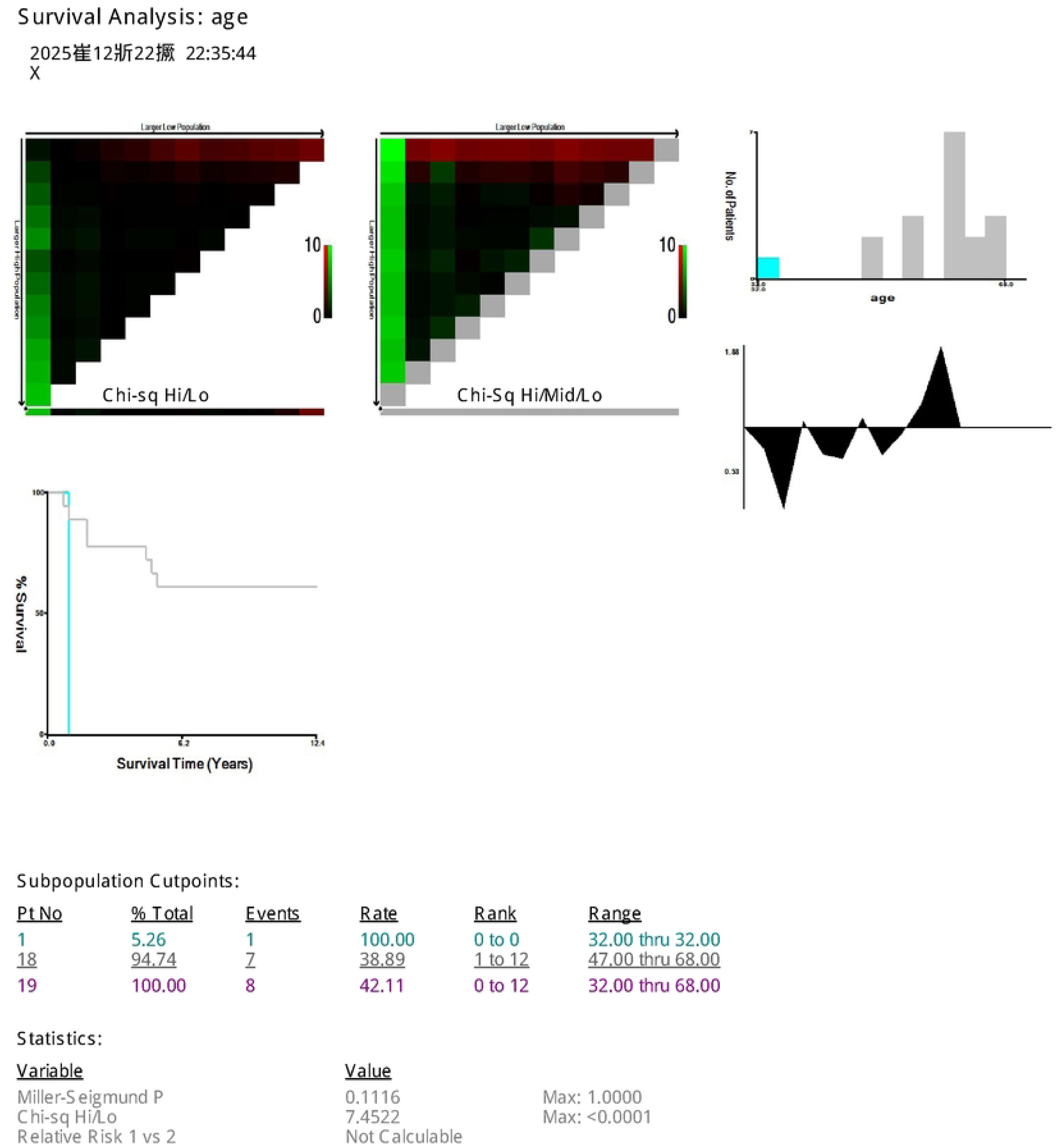
Overall survival curves for two groups after liver transplantation.

**Fig 9.**
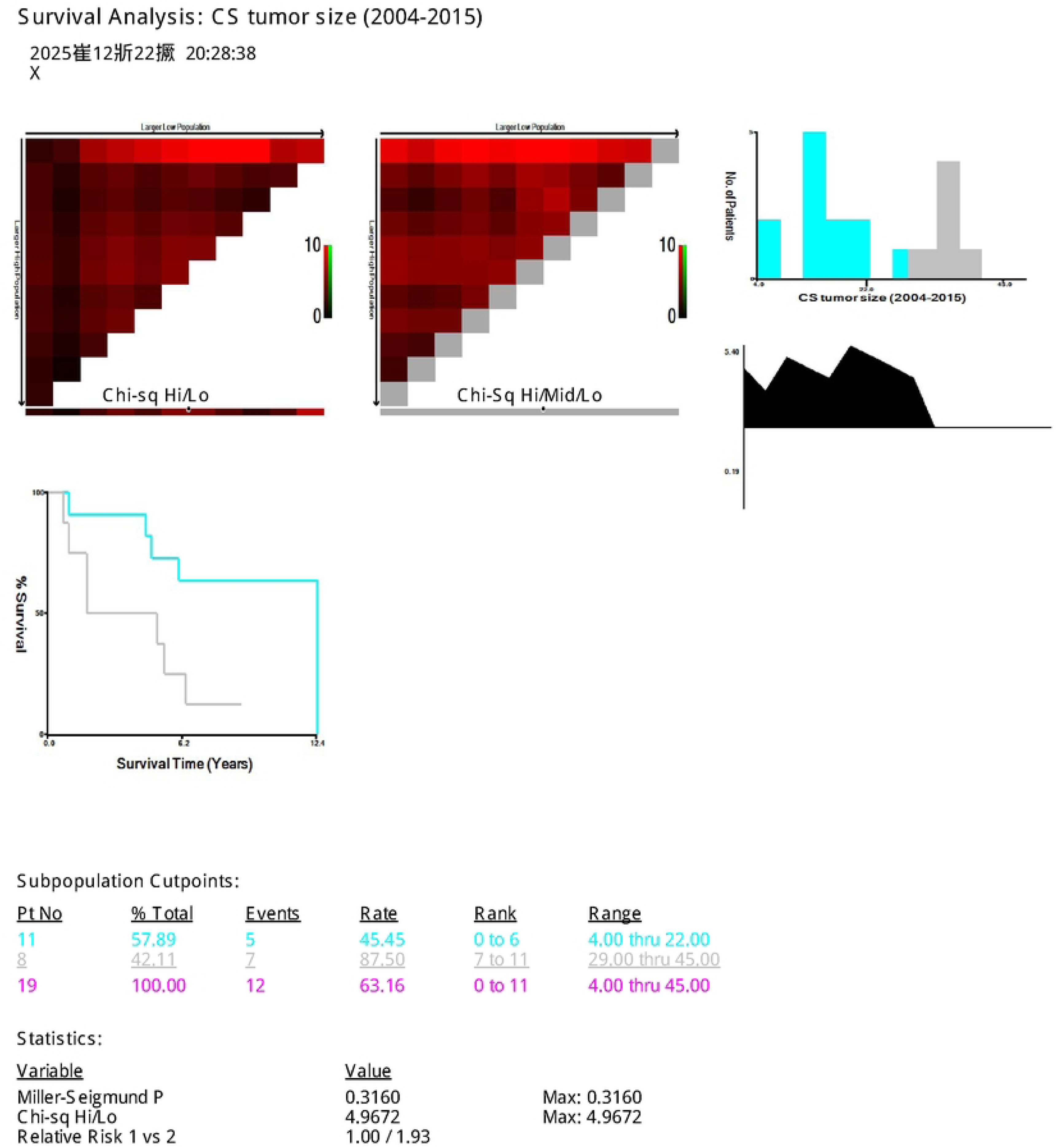
Overall survival curves for two groups after liver transplantation.

**Fig 10.**
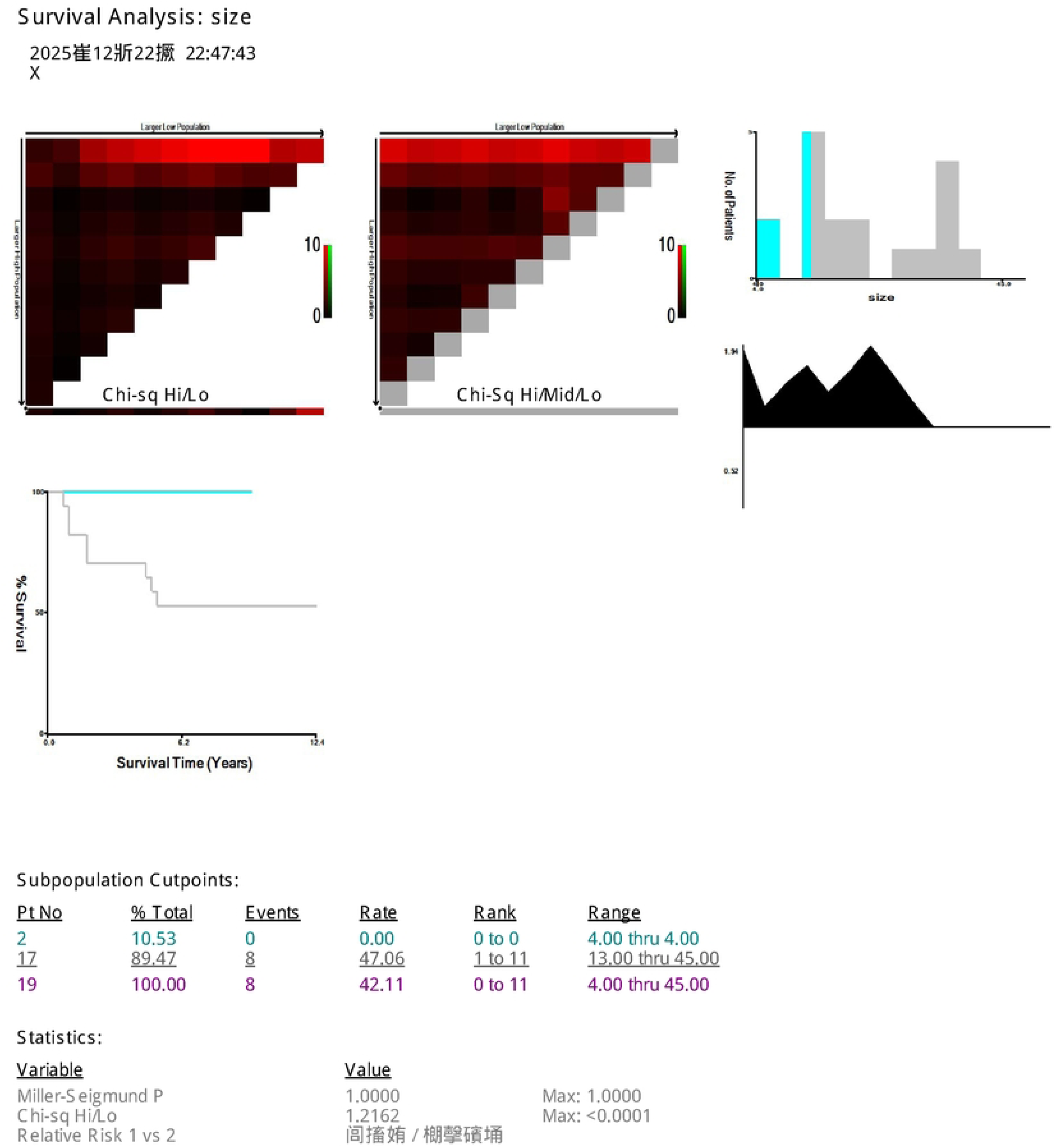
Overall survival curves for two groups after liver transplantation.

**Table 5.** Univariate and Multivariate analyses for Overall survival in ICC patients.

| Variable factors | Overall survival<br>Univariate analysis |  | Multivariate analysis |  |
| --- | --- | --- | --- | --- |
|  | HR (95% CI) | P value | HR (95% CI) | P value |
| Age |  |  |  |  |
| >=60years | Ref |  |  |  |
| <60years | 1.088(0.331-3.576) | 0.889 |  |  |
| Sex |  |  |  |  |
| Female | Ref |  |  |  |
| Male | 1.108(0.324-3.789) | 0.871 |  |  |
| Race |  |  |  |  |
| Other | Ref |  |  |  |
| White | 0.342(0.040-2.933) | 0.328 |  |  |
| Black | 0.975(0.084-11.343) | 0.984 |  |  |
| Marriage |  |  |  |  |
| Unmarried | Ref |  |  |  |
| Married | 1.315(0.348-4.970) | 0.686 |  |  |
| Chemotherapy |  |  |  |  |
| None | Ref |  |  |  |
| Done | 0.901(0.724-1.122) | 0.353 |  |  |
| Radiation |  |  |  |  |
| None | - |  |  |  |

| Variable factors | Overall survival |  | Multivariate analysis |  |
| --- | --- | --- | --- | --- |
|  | Univariate analysis |  |  |  |
|  | HR (95% CI) | P value | HR (95% CI) | P value |
| Done | - | - |  |  |
| Tumor size |  |  |  |  |
| >=29mm | Ref |  |  |  |
| <29mm | 0.265(0.076-0.922) | 0.037 |  |  |
| Lymph node |  |  |  |  |
| >=4piece | Ref |  |  |  |
| None | 21017.908 (0.000-4.599E+153) | 0.955 |  |  |
| 1-3piece | 47289.001<br>(0.000-1.035E+154) | 0.951 |  |  |

Abbreviations: HR, hazard ratio; CI, confidence interval.

**Table 6.** Univariate and Multivariate analyses for Cancer-specific survival in ICC patients.

| Variable factors | Cancer-specific survival |  | Multivariate analysis |  |
| --- | --- | --- | --- | --- |
|  | Univariate analysis |  |  |  |
|  | HR (95% CI) | P value | HR (95% CI) | P value |
| Age |  |  |  |  |
| >=47years | Ref |  |  |  |
| <47years | 8.662(0.785-95.550) | 0.078 |  |  |
| Sex |  |  |  |  |
| Female | Ref |  |  |  |
| Male | 1.029(0.246-4.311) | 0.968 |  |  |
| Race |  |  |  |  |
| Other | Ref |  |  |  |
| White | 0.376(0.044-3.200) | 0.371 |  |  |
| Black | 0.631(0.039-10.342) | 0.747 |  |  |
| Marriage |  |  |  |  |
| Unmarried | Ref |  |  |  |
| Married | 1.391(0.281-6.899) | 0.686 |  |  |
| Chemotherapy |  |  |  |  |
| None | Ref |  |  |  |
| Done | 3.629(0.858-15.356) | 0.080 |  |  |
| Radiation |  |  |  |  |
| None | Ref |  |  |  |
| Done | 1.252(0.399-3.930) | 0.700 |  |  |
| Tumor size |  |  |  |  |
| >=13mm | Ref |  |  |  |
| <13mm | 0.040(0.001-323.602) | 0.483 |  |  |
| Lymph node |  |  |  |  |
| >=4piece | Ref |  |  |  |
| None | 7306.582<br>(0.000-5.596E+116) | 0.947 |  |  |
| 1-3piece | 15039.791<br>(0.000-1.152E+117) | 0.942 |  |  |

Abbreviations: HR, hazard ratio; CI, confidence interval.

## Discussion

This study, based on the SEER database, compared long-term survival outcomes and influencing factors after LT in patients with stage I HCC and ICC according to the 6th edition of the AJCC staging system. Results revealed distinct characteristics between HCC and ICC: ICC patients demonstrated significantly lower 5-year OS and CSS compared to HCC patients. Significant differences in prognostic risk factors were also observed between the two groups. Findings support the critical importance of considering tumor histological type in developing LT strategies and assessing prognosis for early-stage primary liver cancer.

Survival analysis revealed that the 5-year OS and CSS rates in the ICC group were both 82.3%, significantly lower than those in the HCC group (OS: 95.1%; CSS: 97.7%). This finding aligns with most literature reports, potentially attributable to ICC’s more aggressive biological behavior—such as higher tendencies for lymph node metastasis, nerve invasion, and vascular invasion—which substantially increases postoperative recurrence risk[15–17]. Even after rigorous selection, long-term oncological outcomes for ICC remain unfavorable, perpetuating ongoing debate regarding the indications and efficacy of liver transplantation in ICC treatment[18,19].

COX regression analysis further revealed heterogeneity in prognostic factors between the two patient groups. Among HCC patients, age and marital status were independent risk factors for OS, while tum or size was an independent risk factor for CSS. Age and marital status primarily influence long-term survival by affecting patients’ overall health management and support systems: advanced age is a strong predictor of multiple non-tumor-related deaths, and elderly patients face higher risks of postoperative complications and slower recovery[20–22]. Marital status, as a key indicator of social support, correlates with better treatment adherence, psychological well-being, and caregiving, thereby reducing mortality from non-cancer-specific causes[23–25]. Tumor size directly reflects tumor burden and malignant potential, serving as a core biological determinant of recurrence and cancer-specific mortality[26,27]. However, in ICC patients, only tumor size correlates with OS, and no significant risk factors for CS S were identified. This suggests that in ICC, tumor size serves not only as an indicator of local disease but also as a macro-level manifestation of systemic tumor burden and aggressiveness[28]. The inherent high invasiveness of ICC results in a uniformly elevated and relatively homogeneous risk of postoperative recurrence. Conventional clinical-pathological variables struggle to further differentiate cancer-specific mortality risk, suggesting prognosis may depend more on deeper biological contexts such as molecular subtypes and microenvironmental characteristics[29–31]. Thus, fundamental differences exist in the survival determinants between HCC and ICC: HCC patient survival results from the combined effects of combating the cancer itself (dominated by tumor biology) and maintaining overall bodily function (influenced by factors like age and social support). In contrast, ICC is fundamentally closer to a systemic disease. Its treatment strategy must shift from the “screening-local treatment” model used for HCC to a new paradigm centered on systemic therapy with local treatment as adjunctive support. There is an urgent need for bio marker research to achieve prognostic stratification and therapeutic breakthroughs.

This study has certain limitations. First, as a retrospective study, it inevitably carries the risk of patient selection bias. Second, the sample size in the ICC group was too small. While this accurately reflects the low number of actual cases undergoing LT in ICC, it severely limits the reliability of statistical conclusions. Particularly in multivariate analysis, results may exhibit significant uncertainty, and effective subgroup analysis cannot be performed. Finally, the SEER database lacks critical clinical and pathological information, such as vascular invasion, microvascular invasion, and specific neoadjuvant and adjuvant treatment regimens—all important variables influencing HCC prognosis. Despite these limitations, the findings offer clinical insights. For HCC patients, liver transplantation demonstrates excellent long-term survival outcomes. Enhanced postoperative follow-up and management for elderly, unmarried patients and those with larger tumors may help improve prognosis. For ICC patients, extreme caution is warranted when considering liver transplantation as a treatment option, emphasizing stricter preoperative evaluation, particularly regarding tumor size. Future efforts should urgently focus on multicenter collaboration to establish larger ICC liver transplantation cohorts and integrate more comprehensive molecular pathological indicators. This will enable more precise identification of ICC subgroups likely to benefit from liver transplantation and facilitate the development of prognostic prediction models for ICC.

## Conclusions

In summary, LT offers curative potential for HCC patients, but its therapeutic value in ICC remains limited with relatively poor prognosis. Expanding ICC research cohorts and deepening investigations into its molecular biology are crucial for clarifying the role of LT and improving patient outcomes.

## Data Availability

The data underlying the results presented in the study are available from seer.cancer.gov

seer.cancer.gov

